# Estimating the Smallest Worthwhile Difference (SWD) of Psychotherapy for Depression: Protocol for a Cross-Sectional Survey

**DOI:** 10.1101/2024.10.31.24316524

**Authors:** Ethan Sahker, Yan Luo, Kenji Omae, Aran Tajika, Manuela L. Ferreira, Astrid Chevance, Stefan Leucht, Sarah Markham, Roger Ede, Andrea Cipriani, Georgia Salanti, Toshi A. Furukawa

**Author notes:** **Correspondence to:** Ethan Sahker, Population Health and Policy Research Unit, Center for Medical Education and Internationalization, Graduate School of Medicine, Kyoto University, Yoshida-Konoe-cho, Sakyo-ku, Kyoto, 606-8501, Japan.

## Abstract

**Background:** Psychotherapy is proven efficacious for the treatment of depression. However, the patient-perceived importance of its effect is not fully appreciated in the evidence base. The smallest worthwhile difference (SWD) represents the smallest beneficial effect of an intervention that patients deem worthwhile given the harms, expenses, and inconveniences associated with the intervention, and facilitates the interpretation of patient perceived importance of an intervention.

**Methods:** The proposed study will estimate the SWD of the American Psychological Association recommended psychotherapies for depression treatment with English-speaking American and British respondents aged 18 and older. Respondents will be recruited using research participant crowdsourcing sites. The SWD will be estimated using the Benefit-Harm Trade-off Method, presenting survey respondents with variable, hypothetical magnitudes of psychotherapy outcomes to find the smallest acceptable effect over a no treatment alternative. The overall average SWD, and subgroup distributions by participant depression treatment experiences and depression symptomology will be described. Participant characteristics will be included in a regression model to explore associated variability in the SWD.

**Expected Results:** We expect to find an estimate of the SWD for psychotherapy in the treatment of depression. Further, we expect that the SWD will vary between clinical subgroups based on depression symptomology and treatment experiences. We expect there to be variation between those who demonstrate depression symptoms consistent with a depressive disorder, based on PHQ-9 scores or self-reported diagnosis history. We think that the findings from this project will inform the treatment decision process about psychotherapy during the clinical consultation for people with depression.

## INTRODUCTION

Depression is the second leading cause of global disability (GBD 2019 Mental Disorders Collaborators, 2022). Firstline depression treatments listed in most guideline recommendations include either antidepressants or psychotherapy (Cuijpers, Stringaris, et al., 2020). A recent network meta-analysis showed that extended combination therapy is the most efficacious for long-term sustained response, but brief combination and brief psychotherapy have very similar effects and are superior to antidepressants alone (Furukawa et al., 2021). Of course, psychotherapy has significantly fewer risks than antidepressants, which may be more attractive. Yet, psychotherapy can be more expensive for some and may be viewed as more inconvenient. Moreover, many people improve without treatment (natural recovery) (Kendrick et al., 2009; ten Have et al., 2017) and may not believe psychotherapy is worth it, given the associated burdens (i.e., harms, expenses, and inconveniences). When considering the best approach for individualized treatment planning, both the benefits and the burdens need to be considered in relation to the treatment alternatives. Natural recovery and psychotherapy are two options patients consider when weighing the costs and benefits of treatment for depression.

The natural recovery, or no treatment, response rate (50%+ reduction in depression severity) for major depression is estimated between 30-40% after eight weeks (Kendrick et al., 2009; ten Have et al., 2017). The placebo response rate in antidepressant trials is estimated to be 37% (Furukawa et al., 2016). However, placebo may produce a greater response than no treatment due in part to the “placebo effect” (Michopoulos et al., 2021). The true response rate experienced by patients not receiving treatment is difficult to estimate in clinical trials. However, a large pragmatic trial with primary care patients comparing watchful waiting versus antidepressants found the watchful waiting response to be 29% (Kendrick et al., 2009). Therefore, a conservative no treatment response rate would be about 30% (Sahker et al., 2024). The patient-perceived worthwhile effect of antidepressant treatment for depression has been recently estimated to be 55% (IQR: 40 to 75%), when considering potential benefits and burdens (i.e., harms, expenses, and inconveniences) (Sahker et al., 2024). Alternatively, psychotherapy is the other first-line depression treatment option both providers and patients consider but it involves unique benefits and burdens. Yet, the patient-perceived worthiness of psychotherapy has not been estimated.

The psychotherapy response rate for acute phase depression treatment is estimated to be 41% for evidence-based and commonly provided modalities (Cuijpers et al., 2021b). This estimate is derived from largest meta-analysis of psychotherapy trials for depression to date (331 RCTs, 34,285 patients), with all treatment modalities being more efficacious than care as usual (CAU) (Cuijpers et al., 2021b). Cognitive behavioral therapy (CBT) currently comprises the bulk of the up-to-date and robust evidence, demonstrating that patients experience greater treatment response with individual CBT than CAU (OR=2.13), Wait-list (OR=4.00), pill placebo (OR=1.60 to 2.08), and no treatment (OR = 2.07) (Cuijpers et al., 2021; Furukawa et al., 2014). While psychotherapy includes many different modalities for depression treatment, a meta-analysis reported equivalent efficacy across commonly provided modalities (Cuijpers et al., 2008, 2021a). These modalities include behavioral therapy, cognitive therapy, CBT, mindfulness-based cognitive therapy, interpersonal psychotherapy, psychodynamic therapies, and supportive therapy provided in either individual face-to-face, individual online, or group therapy (American Psychological Association, 2019) and can last between 8 and 52 weeks (Cuijpers et al., 2021; Furukawa et al., 2014). These findings, and subsequent guideline recommendations, represent evidence of effectiveness solely determined by researchers. While these determinations are in support of patient treatment outcomes, they do not account for the patient perspective on commonly provided psychotherapies.

The minimal important change (MIC) can help determine the significance of treatment changes in health outcomes. The MIC, (i.e., minimal important difference, minimal important clinical difference), is the smallest post-treatment change in a health outcome perceived as important (Jaeschke et al., 1989). The MIC is used to interpret patient-reported outcome measures (PROMs) (Carrasco-Labra et al., 2021). The MIC for depression scales has been found to be a 6-point reduction on the Beck Depression Inventory-II (Hiroe et al., 2005) and a 7- to 8-point reduction on the Hamilton Depression Rating Scale 17-item (Leucht et al., 2013). The MIC estimate represents intra-individual change pre- to post-treatment (Hiroe et al., 2005; Leucht et al., 2013). However, the MIC is only applicable to one particular outcome measure or instrument (Devji et al., 2020; Ferreira et al., 2012), is not associated with a specific intervention (McNamara et al., 2015), and lacks an account of benefits and burdens relative to a treatment alternative (Ferreira et al., 2012; McNamara et al., 2015). Thus, a more patient-perceived determination of psychotherapy worthiness, given the treatment benefits and burdens is warranted.

The smallest worthwhile difference (SWD) represents a conceptually different method to estimate patient-perceived intervention importance. The SWD is “the smallest beneficial effect of an intervention that justifies the costs, risks, and inconveniences of that intervention over a treatment alternative” (Ferreira et al., 2012). The SWD is a between-treatments assessment that reflects the average patient’s benefit-burden tradeoff when considering two treatment options. The SWD estimate is patient-derived, intervention-specific, and provides an absolute difference between two treatment options (Ferreira et al., 2012; McNamara et al., 2015). There are two accepted methods used for estimating the SWD, the discrete choice experiment and the benefit-harm trade-off method (BHTM). Discrete choice experiments query preferences for hypothetical scenarios at varying levels of benefits and costs, providing a threshold for one treatment over another (Franco et al., 2016). The BHTM queries individual preferences for clinical scenarios varying only the benefits, leaving the burdens static. The BHTM represents the benefits people are willing to accept, given the expected burdens of one intervention over another (Barrett et al., 2005). The BHTM is easy to understand and has been used to estimate the SWD in antidepressant treatments for depression (Sahker et al., 2024). However, the SWD of psychotherapy for depression has never been estimated.

## OBJECTIVE

We have previously estimated the SWD of antidepressant treatment for depression (Sahker et al., 2024), and the present study will extend this research to include patient perspectives on the other major treatment option for depression. The present study will estimate the SWD of psychotherapies recommended by the American Psychological Association for depressive disorders compared to the no treatment/natural course alternative using the BHTM. We will primarily estimate the SWD for the general population with at least moderate depression symptoms who are not receiving any treatment (pharmacological or psychological). As secondary analyses, we will estimate the SWD of those currently in treatment and those without any depressive symptoms. We will also explore demographic and clinical predictors of the SWD. Additionally, we will determine consistency and comparability with the established antidepressant SWD. This study will help mental health providers understand differential patient expectations of psychotherapy and establish an evidence-based benchmark for clinical trials.

## METHODS

### Study design

The present study was approved by the Kyoto University Graduate School of Medicine Ethics Committee (R4578-1), and all participants will provide e-consent. We will conduct a cross-sectional survey using two research participant crowdsourcing services (RPCSs): Prolific and MQ Mental Health (MQ). We will invite participation through RPCSs and link participants fitting the inclusion criteria to an online survey. While Prolific represents a general internet population, MQ includes people with lived experiences in mental health who volunteer to improve research representation. RPCS participants generally demonstrate high test-retest reliability and high convergent and concurrent validity in psychological tests (Chandler & Shapiro, 2016). Prolific participants will be limited to the UK or USA. MQ participants will be limited to the UK only. These RPCSs will provide monetary compensation for participation from their established participant pools. The compensation will be £1.20 GBP (equivalent USD for USA) for Prolific and voluntary for MQ. Remuneration is commensurate with similar length RPCS studies (Sahker et al., 2024) based on time-to-completion. Study information will be provided, and electronic consent will be collected prior to data collection (Appendix 1).

### Primary outcome and its measurement

Our primary outcome is the smallest worthwhile difference (SWD), representing the patient-preferred efficacy dimensions of depression treatment with psychotherapy that would be deemed worthwhile compared to no treatment, given the treatment burdens (harms, expenses, and inconveniences). The measure of efficacy will be treatment response, defined as a 50% or greater reduction in depression symptoms severity eight weeks after psychotherapy initiation. We describe this response to participants as, “feeling much better,” which corresponds with the Clinical Global Impressions Scale and the standard 50% symptom reduction definition of response (Busner & Targum, 2007; Cuijpers, Noma, et al., 2020; Sahker et al., 2024). We define psychotherapy as the American Psychological Association (APA) recommended modalities and formats for treating depression, which includes: behavioral therapy, cognitive therapy, cognitive CBT, mindfulness-based cognitive therapy, interpersonal psychotherapy, psychodynamic therapy, and supportive therapy consisting of individual face-to-face, individual online, or group therapies (American Psychological Association, 2019). We assume that 30 out of 100 people without treatment would experience a minimum 50% reduction in depression symptoms after eight weeks, based on the literature (Toshi A Furukawa et al., 2016; Kendrick et al., 2009; ten Have et al., 2017).

We will estimate the individual participant preference through the Benefit-Harm Tradeoff Method (BHTM) (Figure 1). The BHTM follows four steps: 1) we will present a summary of major depressive episode symptoms, 2) we will explain the benefits of the no treatment alternative to be a 30% response rate after eight weeks, 3) we will explain the potential burdens of APA recommended psychotherapies (American Psychological Association, 2019) in terms of risks (Linden & Schermuly-Haupt, 2014), treatment length (Cuijpers et al., 2013), fees (Benson & Song, 2020; Woods, 2023), and wait times (Baker & Kirk-Wade, 2024; National Council for Mental Wellbeing, 2022), and 4) we will present a series of varying treatment benefits compared to the no treatment alternative of 30% response after eight weeks. In step 4, we will ask participants, “Given the potential drawbacks after 8 weeks, if *x*/100 people going to psychotherapy felt much better (instead of 30/100 from no treatment), would you think the treatment is worthwhile?” (Appendix 2).

**Figure 1.**
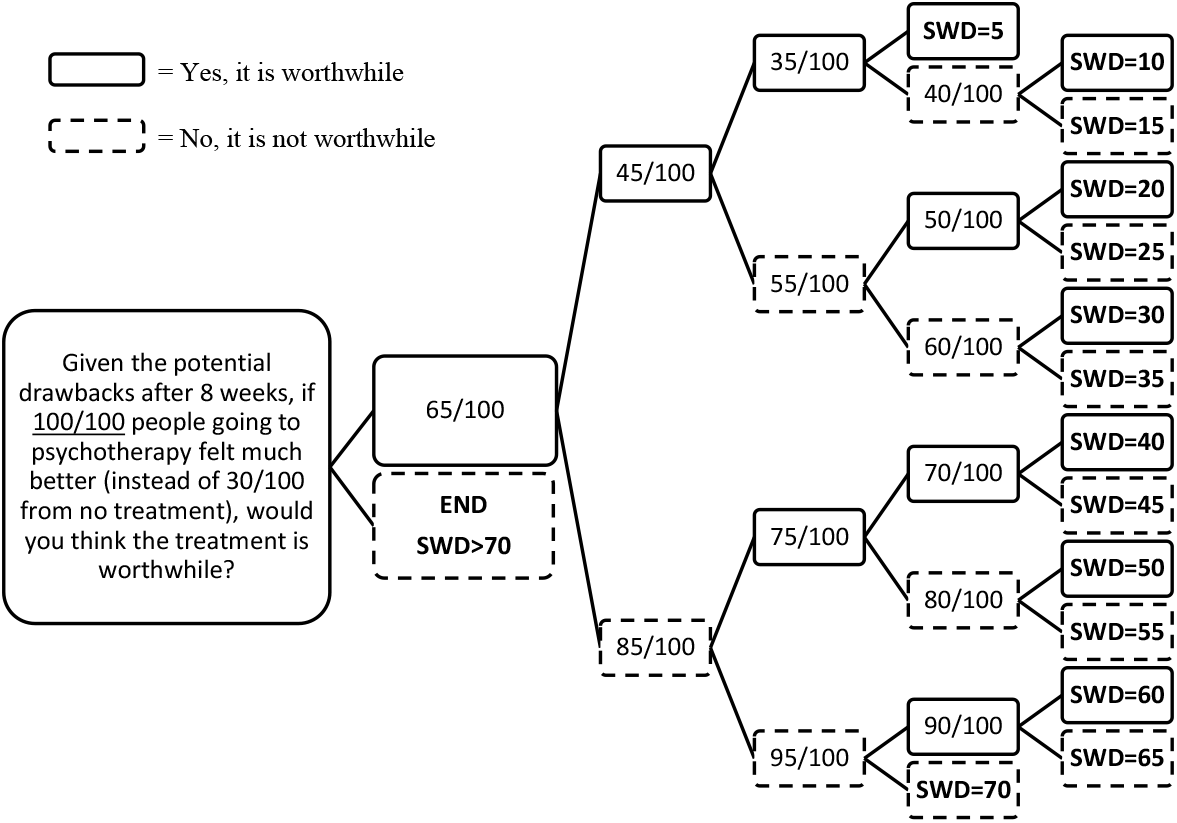
Smallest worthwhile difference (SWD) question sequence algorithm. Participants are queried about their willingness to accept psychotherapy, given the treatment benefits and burdens, until they reach the smallest worthwhile response rate. The first question is repeated but the ratio is replaced by the ratio in the box. The smallest worthwhile response, minus the 30% no treatment response, equals the individual participant SWD.

If participants believe psychotherapy is worthwhile at the variable hypothetical response rate, given the burdens, they will move to a lower response benefit. If they do not believe it is worthwhile, they will move to a higher response rate. The process will continue until the smallest worthwhile response rate is identified. Finally, the difference between the 30% No Treatment response and the smallest psychotherapy response the participant would consider worthwhile represents the individual’s SWD.

Participants meeting the inclusion criteria will be randomly assigned 3:1 to a BHTM for psychotherapy or antidepressants, respectively. We expect this secondary analysis of the antidepressant SWD to replicate the established SWD of antidepressants for depression (Sahker et al., 2024), and to determine the comparability and consistency between antidepressant therapy and psychotherapy for depression.

### Demographic and clinical variables

We will collect demographic information including gender, age, race/ethnicity, education, employment, country of residence (USA, England, Scotland, Wales, and Northern Ireland), and insurance status. Insurance will be categorized as Affordable Care Act (USA only), Medicare/Medicaid (USA only), national healthcare insurance (e.g., NHS, NSE), private health insurance, other, or uninsured.

Clinical variables query participants to affirm or deny lifetime depression diagnosis, family history of depression, lifetime antidepressant treatment, lifetime psychotherapy, current antidepressant treatment, current psychotherapy, and treatment preference (antidepressants vs. psychotherapy). Current depression symptom severity will be also be collected with the Patient Health Questionnaire-9 (PHQ-9) (Kroenke & Spitzer, 2002).

### Participants

We will include participants aged 18 or older, living in the UK or USA. We are interested in a general population and people experiencing depressive symptoms of at least moderate severity (PHQ-9≥10) but not currently engaged in any treatment. This group most closely resembles potential treatment seekers, and we can expect them to provide more accurate estimates of the SWD for a major depressive episode as depicted in the provided clinical scenario because of their current experiences and potential treatment needs. This group would represent the best estimate of a clinical sample taken from a general internet population. To further explore how different experiences with depression and treatment could be associated with treatment-seeking judgements and SWD estimates, we will include participants with four differing profiles, 1) *Moderate-to-severe depressive symptoms but not in treatment*: PHQ-9≥10 and not receiving any treatment – the primary interest group for SWD estimation, 2) *Currently in treatment*: ongoing antidepressant therapy or psychotherapy, 3) *Absent-to-mild depressive symptoms with treatment experiences*: zero to mild depression symptoms (PHQ-9<10), no current treatment, but previous psychotherapy or antidepressant treatment, and 4) *Absent-to-mild depressive symptoms without treatment experiences*: zero to mild depression symptoms and no current or previous psychotherapy or antidepressant treatment.

### Sample Size

The sample size is set to achieve the expected precision in the estimate of the SWD. We assume that the standard deviation (SD) of the SWD would resemble that estimated in our previous study using the same proposed sample with an IQR=10, 35 (approximate SD=20) for SWDs of 20% (Sahker et al., 2024). To obtain a 95% confidence interval (CI) within 10 percentage points, we need approximately 62 participants in each of the four groups described above. Estimates of depression incidence in RPCSs vary but are demonstrably higher than in the general population (Chandler & Shapiro, 2016). Based on our previous distribution (Sahker et al., 2024), we estimate 12% of the sample will comprise the primary interest group *moderate-to-severe depressive symptoms but not in treatment*, 44% currently in treatment, 11% absent-to-mild depressive symptoms with treatment experiences, and 33% absent-to-mild depressive symptoms without treatment experiences. Based on these subgroup population estimates, approximately 518 participants would be necessary to reach n=62 in the groups with the smallest populations (Figure 2). As we will be allocated one quarter of the participants to the antidepressant SWD study, we will recruit approximately 700 participants. Recruitment will be stopped after all four groups include 62 participants, and we will accept responses with no missing outcome variables.

**Figure 2.**
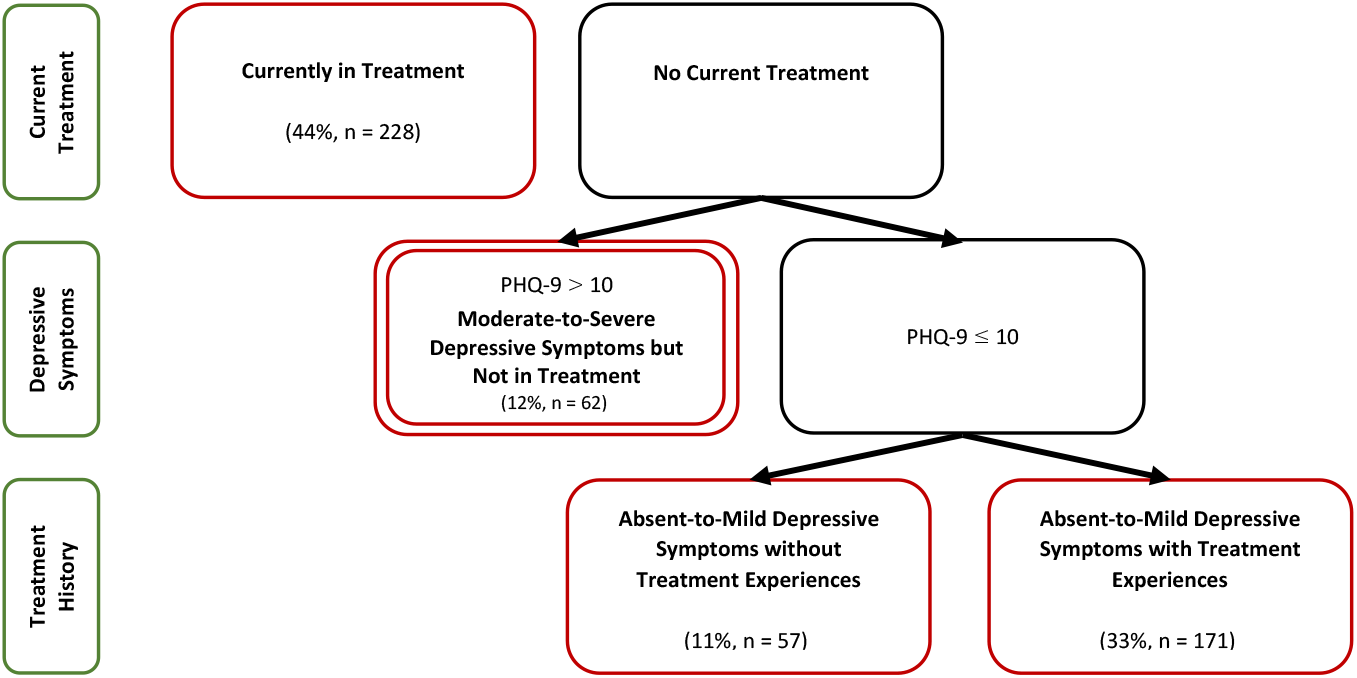
Estimated clinical group representations based on symptomology and treatment experiences. The primary interest being moderate-to-severe depressive symptoms but not in treatment. Red boxes represent subgroups of interest with necessary minimum of n = 62 to calculate the clinical group smallest worthwhile differences, PHQ-9 = Personal Health Questionnaire 9-item, % = estimated from population.

### Data Analysis

We will first present the SWD with its distribution and percentile ranking for people with moderate-to-severe depressive symptoms but not in treatment and estimate its median and interquartile range (IQR). We will then examine the SWD by participant groups for comparison using box-violin plots. Finally, we will use exploratory methods to analyze the entire sample’s SWD using demographic and clinical independent variables in univariable regressions and a single multivariable regression investigating SWD predictors using the least absolute shrinkage and selection operator (LASSO) method (Tibshirani, 1996). We will only accept responses with no missing outcome variable data. Participants who report they would not accept psychotherapy, even if the response were 100%, will be removed from the primary analysis because they would not be real-world candidates for the treatment (McNamara et al., 2015; Sahker et al., 2024). To examine the effect of this decision, we will conduct sensitivity analyses by assigning them an SWD value representing a psychotherapy response rate over 100% (SWD=71). We will use SAS 9.4 (Cary, NC, SAS Institute Inc) and R 4.2.2 (R Core Team, 2022) for all statistical analyses.

### Patient and public involvement

We will pilot the BHTM script with two members of the Patient and Public Involvement (PPI) group at the Oxford Precision Psychiatry Lab of the University of Oxford, who are also co-authors of this project (SM and DE). PPI members will review descriptions of a DSM-5 major depressive episode, and the benefits and burdens of psychotherapy. These experts by experience will provide feedback on clarity, inclusivity, and accuracy of patient experiences, and we will modify the scripts accordingly. They will further collaborate in the interpretation and write-up of the manuscript.

### Expected Results

We expect to find an estimate of the SWD for psychotherapy in the treatment of depression for the first time. Further, we expect that the SWD will vary between clinical and demographic subgroups. We expect there to be variation between those who demonstrate depression symptoms consistent with a depressive disorder, based on PHQ-9 scores or self-reported diagnosis history. In addition, we believe that the present study’s results for the antidepressants SWD will be consistent with previous findings, and this would strengthen the comparability of psychotherapy and antidepressant SWDs. We think that the findings from this project will inform clinical practices in treatment consultation and decision-making for psychotherapy for depression.

## Data Availability

All data produced in the present study are available upon reasonable request to the authors.

## ACKNOWLEDGMENTS

ES is supported by the Japan Society for the Promotion of Science (grant 24K20239). ACi is supported by the National Institute for Health Research (NIHR) Oxford Cognitive Health Clinical Research Facility, by an NIHR Research Professorship (grant RP-2017-08-ST2-006), by the NIHR Oxford and Thames Valley Applied Research Collaboration and by the NIHR Oxford Health Biomedical Research Centre (grant BRC-1215-20005). The views expressed are those of the authors and not necessarily those of the UK National Health Service, the NIHR, or the UK Department of Health. MLF holds a National Health and Medical Research Council of Australia Investigator Fellowship.

## ETHICAL APPROVAL

The present study was approved by the Kyoto University Graduate School of Medicine Ethics Committee (R3574-1), and all participants provided e-consent.

