## Appendix for "Estimating the Smallest Worthwhile Difference (SWD) of Psychotherapy for Depression: Protocol for a Cross-Sectional Survey"

**Appendix 1.** Informed e-Consent Information Statement

**Appendix 2.** Smallest Worthwhile Difference Survey Scenario for the Benefit Harm Tradeoff Method (BHTM)

**Appendix 1.** Informed e-Consent Information Statement

**Electronic Description Document**

**Study Title:** Estimating the Smallest Worthwhile Difference (SWD) of Psychotherapy for Major Depressive Disorder

**Research Summary**:

***Ethics review and permission***: Prior to onset, this study was reviewed and approved by the Medical Ethics Committee of the Kyoto University Graduate School of Medicine and Faculty of Medicine, the Kyoto University Hospital, and the head of the research institute.

***Name of research institute and name of principal investigator***: We are researchers at Kyoto University, led by Dr. Ethan Sahker, PhD, in collaboration internationally with researchers at the University of Bern (Professor Georgia Salanti, PhD), University of Oxford (Professor Andrea Cipriani, MD, PhD), and the University of Sydney (Professor Manuela L Ferreira, PhD).

***Reason for being selected as a research subject***: You were selected for this survey because you are representative of the general population aged 18 or older.

***Study objectives and significance***: We are studying patient preferences in the treatment of depression. The purpose of this study is to determine how to evaluate the effectiveness of psychotherapy from the patient's perspective, considering the benefits and disadvantages of psychotherapy. We believe that the survey results will help healthcare professionals and researchers understand patient expectations regarding the effectiveness of psychotherapy and accurately interpret the importance of the results obtained from clinical research.

***Study method***: We plan to recruit approximately 1,000 participants. Those who agree to participate will be asked to complete an online survey that takes approximately 5 to 10 minutes. First, participants will be asked about their mental health and past treatment history. Next, we will explain depression and the effectiveness of psychotherapy, as well as the burdens and costs. We will then ask your opinions on the effectiveness of psychotherapy through a series of questions.

***Duration of the study***: From the date of approval by the head of the research institution to September 30, 2027.

**Risks and Benefits**: Risks to participants are expected to be minimal. There is a time commitment to complete the questionnaire. In addition, collecting data over the Internet and normal Internet use are thought to be equally strenuous. There is no financial burden on participants. Participation in this study may not directly benefit participants, but it provides an opportunity to gain knowledge about depression, as well as the costs and benefits of psychotherapy. It will also contribute to important issues such as how results obtained in future clinical studies could be interpreted to incorporate patient values into clinical trials.

**Financial burden and compensation for research subjects:** In principle, there is no financial burden associated with participating in this study, but participants will be responsible for communication costs associated with connecting to the Internet when answering the questionnaire. Participants will be compensated for the time and effort spent answering the questionnaire in accordance with the terms of each crowdsourcing service.

**Handling of personal information:** Survey participant crowdsourcing website IDs will be managed in accordance with the websites’ Participant Agreement and Privacy Policy. Only the principal investigator will have access to the website ID, but it will be used to allocate rewards and will not be shared with others. Survey data will be stored on the secure servers of the crowdsourcing websites, but they will not have access to survey data. Data will be backed up by the principal investigator. After data collection is complete, all IDs will be deleted, and data will be deidentified to make it unidentifiable. Once the survey is complete, all data will be deleted from websites’ servers. Deidentified data will be stored on a password-protected local server in the principal investigator's lockable room for 10 years after publication of the primary results. Thereafter, the data stored so that personal information cannot be known and identifying information will be destroyed. No personally identifiable information will be included in any papers or conference presentations related to the results of this study. Secondary use in other research and provision to other research institutions will not include any personally identifiable information.

**When providing information to a person in a foreign country:**

1. Universities: University of Bern (Switzerland), University of Oxford (UK), and the University of Sydney (Australia)
2. Information on the system for protecting personal information in the foreign country obtained in an appropriate and reasonable manner: Detail: https://www.ppc.go.jp/personalinfo/legal/kaiseihogohou/#gaikoku
a) University of Bern (Switzerland) - The Federal Act on Data Protection of 19 June 1992 and The Ordinance to the Federal Act on Data Protection of 14 June 1993
   - https://www.fedlex.admin.ch/eli/cc/1993/1945_1945_1945/en
   - https://www.fedlex.admin.ch/eli/cc/1993/1962_1962_1962/en

b)	University of Oxford (UK) - Compatible with the Personal Information Protection Commission of Japan

c)	The University of Sydney (Australia) – Asia-Pacific Economic Cooperation (APEC) Privacy Framework, and an enforcement agency has the authority to investigate and correct complaints that cannot be resolved by Cross Border Privacy Rules (CBPR)-certified businesses or accountability agents. Japan follows the APEC Privacy Framework enforcement and public grievances are handled at the same level. Personal information can be expected to be protected at the same level as in Japan.

- Privacy Act 1988: https://www.legislation.gov.au/Details/C2021C00139
- Privacy Regulation 2013: https://www.legislation.gov.au/Details/F2021C00274

3. Information on the measures taken by the person to protect personal information *See Ethical Guidelines, pages 99-104: ID management for Prolific and MQ Health follows the site's privacy policy. Only the Principal Investigator has access to the participant ID, but it will not be shared with anyone other than the Principal Investigator and will be used only for the purpose of distributing rewards. Survey data will be stored on the secure servers of Prolific and MQ Health, but these companies will not have access to the survey responses. Once data collection is complete, participant IDs will be removed from the dataset and the data will be anonymized so that individuals cannot be identified. The anonymized data will be stored in the principal investigator's locked room on a password-protected local server for 10 years after the publication of the main results. The data will then be erased so that personal information cannot be identified, and the media will be disposed of. The anonymized data will be shared with collaborating universities, but no information will be linked to participants. Reports and presentations on the findings of this study will not contain any personally identifiable information.

**Potential for secondary use of information or provision to other research institutions:** Information collected in this study may be used for future research that is not specified at the time of obtaining consent. When used for secondary research or provided to other research institutions, no personally identifiable information will be included. If you would like to confirm future research, you can contact the principal investigator (see contact information below).

**Voluntary Participation**: Participation in this study is voluntary. Participants may withdraw consent at any time without giving a reason if they maintain their own personally created survey ID. Those withdrawing will not be disadvantaged in any way if they do not agree to participate or if consent is withdrawn. If consent is withdrawn, we will not use the data collected. However, once the analysis results are published, it is difficult to delete the data. Materials related to this research may be obtained and viewed to the extent that it does not interfere with the personal information and intellectual property of other research subjects and others. The decision to participate or not to participate will not affect current or future relationships with Kyoto University or the crowdsourcing websites.

**Research Funding and Conflicts of Interest**: This research is funded by the Kyoto University Graduate School of Medicine. Conflicts of interest are appropriately reviewed by the Kyoto University Clinical Research Conflict of Interest Review Committee in accordance with the Kyoto University Conflict of Interest Policy and the Kyoto University Conflict of Interest Management Regulations.

**Contact Information**: For further information, please contact Dr. Ethan Sahker, Junior Associate Professor, Population Health and Policy Research Unit, Center for Medical Education and Internationalization, Graduate School of Medicine, Kyoto University:.

Kyoto University Contact: Research Promotion Division, General Affairs and Planning Department, Graduate School of Medicine, Kyoto University

**Consent**:

By selecting "I agree" to the first question on the survey form – "Have you read the instructions and agree to participate in this study?" – you agree to participate in this study. Please also read your crowdsourcing website’s Participant Agreement and Privacy Agreement as they pertain to the management of individual website IDs.

**Appendix 2.** Smallest Worthwhile Difference Survey Scenario for the Benefit Harm Tradeoff Method (BHTM)

**[Page 1] -------------------------------**

**The next five slides include information on depression and its treatment. Please carefully read them and keep this in mind when answering the questions after the Information.**

**CLINICAL DEPRESSION**

Everyone experiences “depressed” from time to time. However, clinical depression is a more serious condition lasting much longer. When clinical depression is moderate to severe, symptoms such as the following are present nearly every day for a minimum of two weeks, and often for several months:

- Feelings of sadness, tearfulness, emptiness, or hopelessness
- Loss of interest or pleasure in most or all normal activities, such as hobbies, sports or sex,
- Reduced appetite and weight loss or conversely, increased cravings for food and weight gain
- Anxiety, agitation, or restlessness
- Sleep disturbances, including insomnia or sleeping too much
- Slowed thinking, speaking, or body movements
- Tiredness and lack of energy, so even small tasks may require extra effort
- Feelings of worthlessness or guilt, fixating on past failures or self-blame
- Trouble thinking, concentrating, making decisions, and remembering things
- Frequent or recurring thoughts of death, suicidal thoughts, or suicide attempts
- Unexplained physical problems, such as back pain or headaches

Not everyone who is depressed experiences every symptom. Some people experience only a few symptoms while others may experience many. Symptoms are usually severe enough to cause noticeable problems with relationships, work, school, at social activities.

**[Page 2] -------------------------------**

**Please carefully read the following and keep this in mind when answering the remaining questions.**

**TREATMENT OF CLINICAL DEPRESSION**
There are many different approaches to treating depression, but in this study, we will only focus on two options, (1) no treatment (i.e., natural recovery) and (2) psychotherapy (i.e., psychological intervention, counseling, talk therapy). First, I will describe each treatment option's expected benefits and drawbacks. Then, I will ask if you think psychotherapy is worthwhile at different levels of patient benefit.

**[Page 3] -------------------------------**

**Please carefully read the following and keep this in mind when answering the remaining questions.**

**TREATMENT OF CLINICAL DEPRESSION**

(1) **No Treatment (Natural Recovery)**

By declining all treatment, about 30/100 people can expect to feel much better after 8 weeks on average.

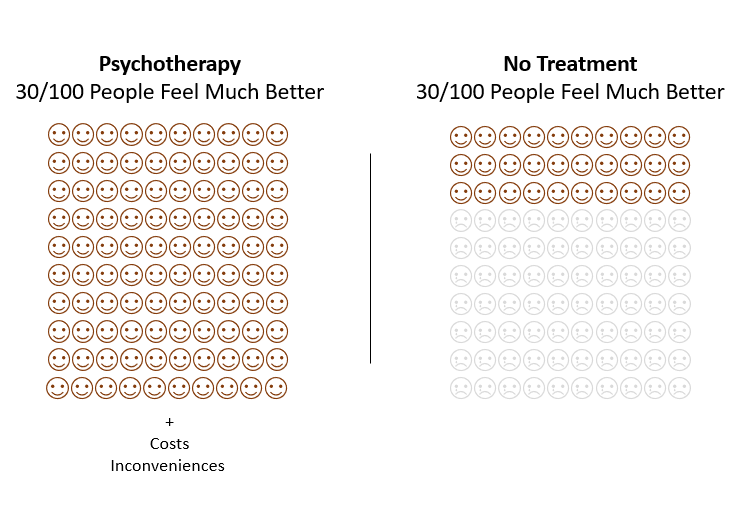

**[Page 4] -------------------------------**

**Please carefully read the following and keep this in mind when answering the remaining questions.**

**TREATMENT OF CLINICAL DEPRESSION**

(2) **Psychotherapy (i.e., psychological intervention, counseling, talk therapy)**

Psychotherapy includes a range of counseling or psychological interventions used to treat clinical depression. There are many different types of commonly administered psychotherapies for depression backed by evidence which are equally effective.

Typically, a course of psychotherapy requires around 11 weekly sessions, each lasting 50 minutes. In these sessions, patients talk with therapists, discuss their problems, gain insight, and find solutions. Psychotherapy usually requires some additional work outside the sessions, called “homework.”

There are no physical side effects associated with psychotherapy, but therapy can bring about unwanted thoughts or behavior changes that can affect one’s relationships and lifestyle. Additionally, there are other burdens that patients should consider, such as scheduling difficulties, treatment costs, travel expenses, time necessary for the visits, and time to be spent on homework.

**[logical Skip if in US]**

**In the UK,** mental healthcare is covered by the National Health Service (NHS), but some people buy supplemental health coverage or pay out of pocket to avoid wait times. Supplemental coverage can cost £50-£150 per session out of pocket. The average wait time for the first treatment is 21 days, and 50 days between the first and second session. Wait times vary considerably based on location, from 4 to 229 days.

**[logical Skip if in UK]**

**In the USA,** a typical 50-minute therapy session can cost between $100-$200, and the average insured session costs $21. The average wait for treatment is 48 days, but this varies widely from same-day services in some areas to long-term waits in rural areas.

**[Page 5] -------------------------------**

**Please carefully read the following and keep this in mind when answering the remaining questions.**

There are many different approaches to treating depression, but please assume you only have two choices when deciding to relieve depressive symptoms: Psychotherapy (i.e., psychological intervention, counseling, talk therapy) versus no treatment at all. Do not consider any other alternatives in your decision.

First, consider the possible drawbacks we’ve mentioned (expenses and other inconveniences). Then, weigh the drawbacks with the presented hypothetical benefits when answering the following questions.

**TRADEOFF QUESTIONS**

**Given the potential drawbacks after 8 weeks, if 100/100 people going to psychotherapy felt much better (instead of 30/100 from no treatment), would you think the treatment is worthwhile?**

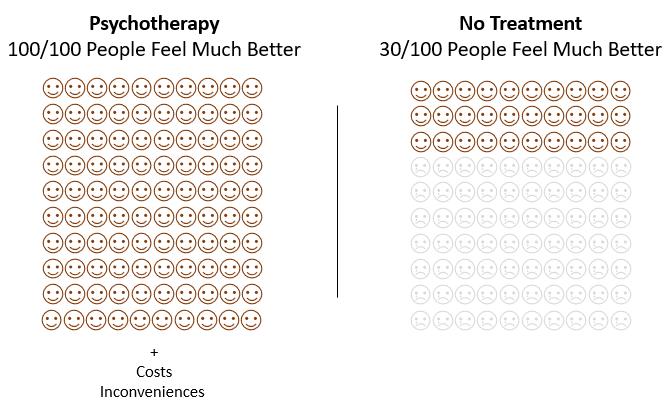

**Respondent responses follow an algorithm presented in Figure 1, altering the response proportion/ratio and the representative psychotherapy face figure.**
